# Two-Stage Adaptive Pooling with RT-qPCR for COVID-19 Screening

**DOI:** 10.1101/2020.07.05.20146936

**Authors:** Anoosheh Heidarzadeh, Krishna Narayanan

## Abstract

We propose two-stage adaptive pooling schemes, 2-STAP and 2-STAMP, for detecting COVID-19 using real-time reverse transcription quantitative polymerase chain reaction (RT-qPCR) test kits. Similar to the Tapestry scheme of Ghosh *et al*., the proposed schemes leverage soft information from the RT-qPCR process about the total viral load in the pool. This is in contrast to conventional group testing schemes where the measurements are Boolean. The proposed schemes provide higher testing throughput than the popularly used Dorfman’s scheme. They also provide higher testing throughput, sensitivity and specificity than the state-of-the-art non-adaptive Tapestry scheme. The number of pipetting operations is lower than state-of-the-art non-adaptive pooling schemes, and is higher than that for the Dorfman’s scheme. The proposed schemes can work with substantially smaller group sizes than non-adaptive schemes and are simple to describe. Monte-Carlo simulations using the statistical model in the work of Ghosh *et al*. (Tapestry) show that 10 infected people in a population of size 961 can be identified with 70.86 tests on the average with a sensitivity of 99.50% and specificity of 99.62%. This is 13.5x, 4.24x, and 1.3x the testing throughput of individual testing, Dorfman’s testing, and the Tapestry scheme, respectively.

## I. Introduction

There is broad consensus among epidemiologists, economists and policy makers that wide-scale testing of asymptomatic patients is the key for reopening the economy. While the benefits of testing are obvious, shortage of testing kits, reagents and the ensuing low-throughput of individual testing protocols has prevented deployment of wide-scale testing. Group testing or, pooling is an alternative way to substantially increase the testing throughput.

The idea of group testing was introduced by Dorfman [1] during World War II for testing soldiers for syphilis without having to test each soldier individually. Dorfman’s scheme consists of two stages (or rounds). In the first stage, the set of people to be tested is split into disjoint pools and a test is performed on each pool. If a pool tested negative, everyone in that pool will be identified as non-infected. Otherwise, if a pool tested positive, we proceed to the second stage where all people in a positive pool will be tested individually, and then identified as infected or non-infected accordingly. When the prevalence is small, Dorfman’s scheme requires substantially fewer tests than individual testing.

Dorfman-style testing has been implemented in the past in screening for many diseases including HIV [2], Chlamydia and Gonorrhea [3]. It has also been considered for screening for influenza [4]. For COVID-19, several experimental results have confirmed the feasibility of using Dorfman-style pooling and it has been implemented in Nebraska in USA, Germany, India and China [5], [6], [7], [8].

While Dorfman-style pooling is easy to implement, it is not optimal. Over the past 75 years, more sophisticated group testing schemes that provide higher testing throughput have been designed. The literature on group testing is too vast to review in detail and an overview of the techniques can be found in [9] and [10]. Group testing is also related to compressed sensing and insights from compressed sensing have been used to design group testing schemes. An important difference between group testing and compressed sensing is that in group testing, the measurements are Boolean (test result is either positive or negative) and they naturally correspond to *non-linear* functions of the unknown vector.

Most of the works using group testing with real-time reverse transcription quantitative polymerase chain reaction (RT-qPCR) have only considered Boolean measurements even though the RT-qPCR process can produce more fine-grained information (soft information) about the total viral load in the pool. It is well-known in information theory that such soft information can potentially be used to increase testing throughput substantially. However, group testing schemes that leverage soft information from the RT-qPCR process remain largely unexplored.

Very recently, Ghosh *et al*. in [11] developed a statistical model relating the soft information from the RT-qPCR to the total viral load in the pool. They designed a scheme called Tapestry, which uses *non-adaptive* group testing using Kirkman triples and they considered several decoding algorithms that use the soft information. They showed substantial gains in testing throughput over Dorfman’s scheme and to the best of our knowledge, this scheme is the state of the art non-adaptive group testing scheme that works with RT-qPCR, especially since it is the only work we are aware of that uses the soft information from the RT-qPCR measurement process.

Here, we propose two simple and effective *two-stage adaptive pooling* schemes that use the soft information from the RT-qPCR process and provide several advantages over Dorfman’s scheme and the Tapestry scheme. We refer to these algorithms as the Two-stage Adaptive Pooling (2-STAP) and the Two-stage Adaptive Mixed Pooling (2-STAMP) schemes/algorithms. The proposed schemes provide substantially higher throughput than Dorfman-style testing. Compared to the Tapestry scheme in [11], 2-STAP and 2-STAMP have higher testing throughput and under the statistical model developed in [11], for all tested cases, our algorithms have higher sensitivity and higher specificity. The proposed algorithms require fewer pipetting operations than Tapestry, but require more pipetting operations than Dorfman’s scheme. Finally, 2-STAP and 2-STAMP work with much smaller pool sizes and population sizes than the Tapestry algorithm and hence, is easy to describe and implement in the lab. Monte-Carlo simulations using the statistical model in the work of Ghosh *et al*. (Tapestry), show that 10 infected people in a population of size 961 can be identified with 70.86 tests on the average with a sensitivity of 99.50% and specificity of 99.62% with a pool size of 31. This is 13.5x, 4.24x, and 1.3x the testing throughput of individual testing, Dorfman’s testing, and the Tapestry scheme, respectively.

Unlike Tapestry, which is a non-adaptive scheme, 2-STAP and 2-STAMP require storage of the swab samples and their accessibility for the second round of testing—similar to that of Dorfman’s scheme.

## II. Measurement Model

We consider pooling-based testing for COVID-19 using the real-time reverse transcription quantitative polymerase chain reaction (RT-qPCR) technique—considered also in [11].

Consider a population of *n* people, labeled 1, …, *n*, that are to be tested for COVID-19. Let *x*_1_, …, *x*_*n*_ represent the viral loads of these people. If the *j*th person is infected (i.e., COVID-19 positive), then *x*_*j*_ is a nonzero value; otherwise, if the *j*th person is not infected (i.e., COVID-19 negative), then *x*_*j*_ is zero. We wish to find the label set of all infected people in the population (e.g., if the people labeled 10 and 20 are the only infected people in the population, the label set of infected people is *{*10, 20*}*), by making measurements using the RT-qPCR technology. A measurement *y* is defined as the sum of the viral loads of a subset (or a pool) of people in the population to be tested for COVID-19. That is,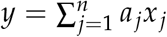, where *a*_*j*_ ∈ *{*0, 1*}*. Suppose we collect the measurements *y*_1_, *y*_2_, … (for different pools), and observe noisy versions of *y*_1_, *y*_2_, …, denoted by *z*_1_, *z*_2_, …. Using the same model as in [11], we assume that the *i* th noisy measurement is given by

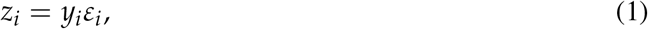

where *ε*_*i*_’s represent the random noise in the measurement process. Note that *z*_*i*_ = 0 if and only if *y*_*i*_ = 0 (i.e., all people participating in the *i*th measurement are non-infected), and *z*_*i*_ ≠ 0 if and only if *y*_*i*_ ≠ 0 (i.e., there exists at least one infected person among the people participating in the *i*th measurement). A detailed explanation about the multiplicative noise model in (1) can be found in Appendix I.

We refer to the process of generating the measurements as *pooling*, and refer to the process of estimating the set of infected people from the noisy measurements as *recovery*. Given a pooling algorithm and a recovery algorithm, the average fraction of infected people that have been identified as non-infected is referred to as the *false negative rate* (denoted by 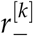), and the average fraction of non-infected people that have been identified as infected is referred to as the *false positive rate* (denoted by 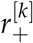), where both averages are taken over all populations of size *n* that contain *k* infected people. The quantities 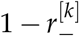 and 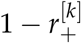 are referred to as the *conditional sensitivity* and the *conditional specificity*, respectively. (Here, the term “conditional” reflects that the false negative rate and the false positive rate are computed by averaging over all populations with a fixed number of infected people.)

Our goal is to design a pooling algorithm and a recovery algorithm such that the total number of measurements is minimized while the false negative rate and the false positive rate remain below some target thresholds (e.g., below 1%), or equivalently, the conditional sensitivity and the conditional specificity remain above some target thresholds (e.g., above 99%).

## III. 2-STAP: A Two-Stage Adaptive Pooling

In this section, we propose a two-stage adaptive pooling scheme which we refer to as the 2-STAP scheme. The 2-STAP scheme is inspired by the well-known two-stage Dorfman’s scheme which was originally proposed in the context of group testing but requires substantially fewer measurements.

Translating Dorfman’s scheme into the language of our work, in the first stage, the people are pooled into a number of disjoint groups (or pools) of equal size, and one measurement is made for each pool where the measurement is the sum of viral loads of all people in that pool. In the second stage of Dorfman’s scheme, one measurement is made for every person in a *positive pool* (i.e., a pool whose measurement in the first stage is nonzero), and no additional measurements are made for any *negative pool* (i.e., a pool whose measurement in the first stage is zero). A schematic of Dorfman’s scheme is shown in Fig. 1.

**Fig. 1.**
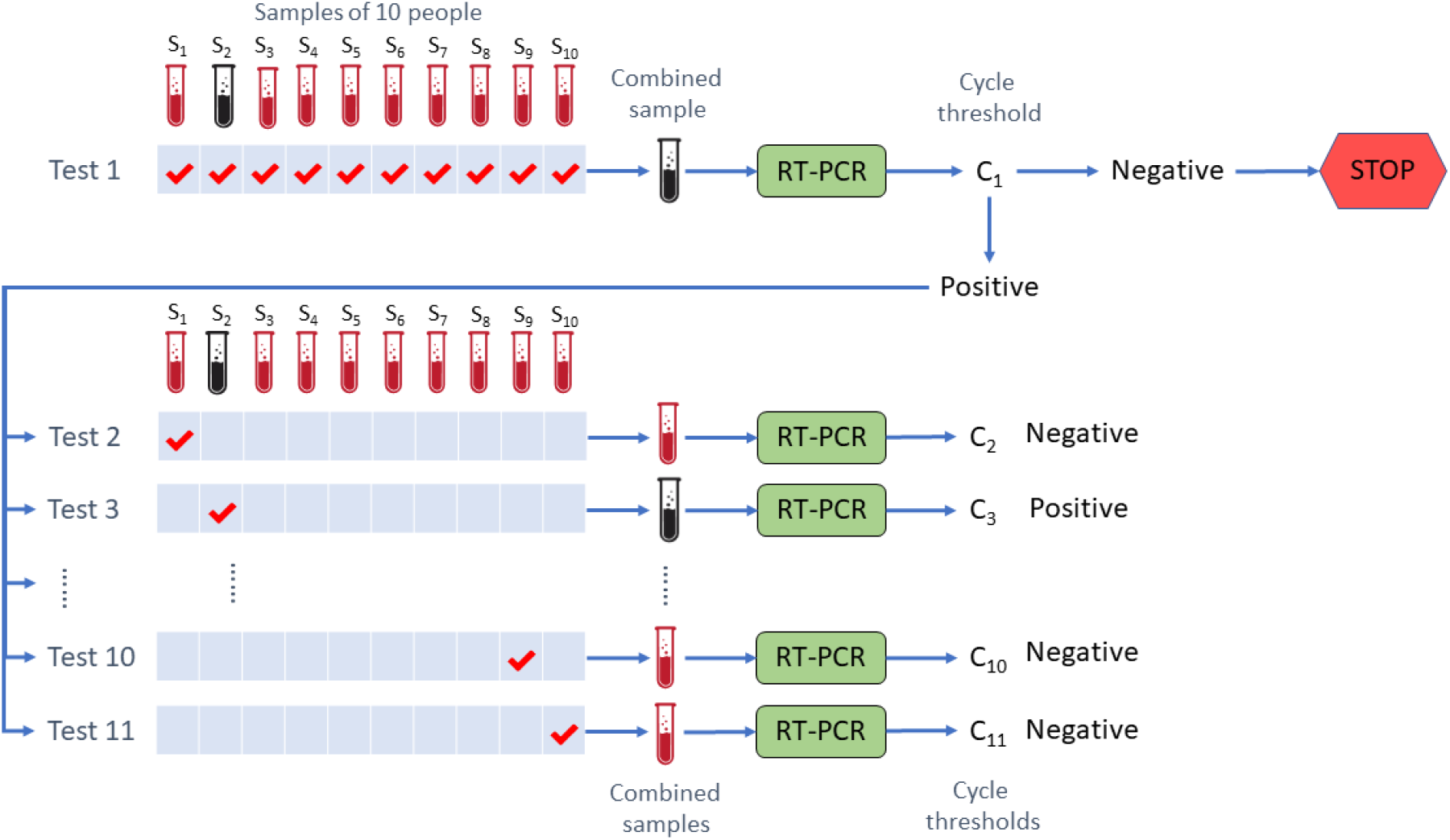
Schematic of Dorfman’s Scheme for pool size = 10.

The first stage of the 2-STAP scheme is the same as that of Dorfman’s scheme. The second stage of the 2-STAP scheme, however, differs from that of Dorfman’s scheme. In particular, in the second stage of the 2-STAP scheme, for each positive pool a number of measurements are made, where each measurement is the sum of viral loads of some (not necessarily singleton) subset of people in that pool. Similar to the Dorfman’s scheme, the second stage of the 2-STAP scheme makes no additional measurements for any negative pools.

We present two variants of the 2-STAP scheme: 2-STAP-I and 2-STAP-II. In 2-STAP-I, for all positive pools, the number of measurements and the pooling scheme in the second stage will be the same, regardless of the observed measurements for these pools in the first stage. On the other hand, in 2-STAP-II, for each positive pool, the number of measurements and the pooling scheme in the second stage will be chosen based on the observed measurement for that pool in the first stage. Although the 2-STAP-II scheme requires a smaller average number of measurements (for a given target level of accuracy), but the 2-STAP-I scheme makes it easier to understand the main advantages of the proposed schemes over non-adaptive schemes and Dorfman’s scheme. A schematic of the 2-STAP-I scheme is shown in Fig. 2.

**Fig. 2.**
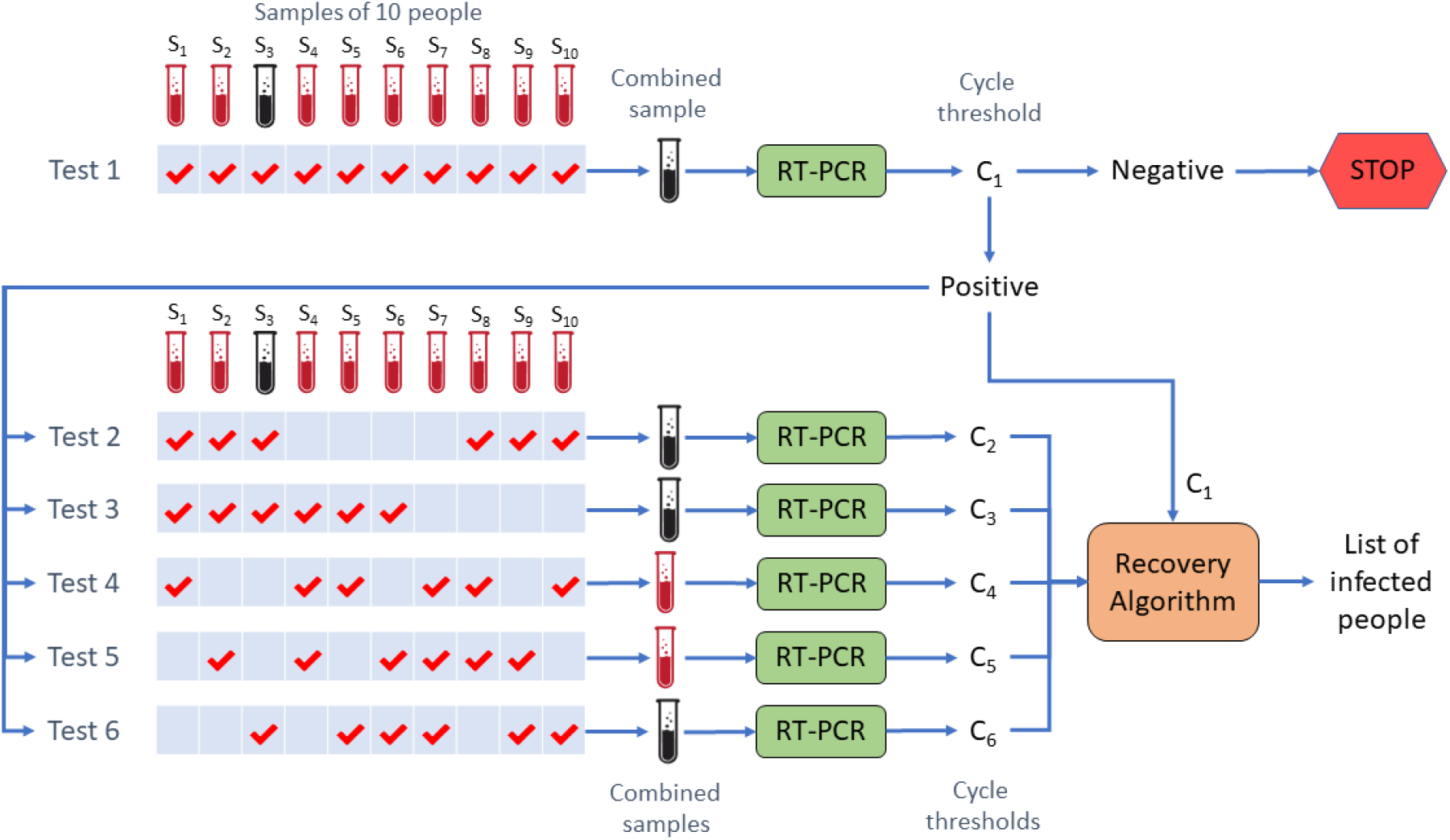
Schematic of 2-STAP-I for pool size = 10.

Once the second-stage measurements for a positive pool are collected, the recovery algorithm takes all these measurements (together with the first-stage measurement for that pool) as input, and returns an estimate of the label set of infected people in that pool. Details about the pooling matrices we have used in our Monte-Carlo simulations can be found in Appendix II, and the detailed mathematical description of the recovery algorithm can be found in [12].

## IV. 2-STAMP: A Two-Stage Adaptive Mixed Pooling

In this section, we propose another two-stage adaptive pooling scheme, which we refer to as the 2-STAMP scheme, by generalizing the 2-STAP scheme. The first stage of the 2-STAMP scheme is the same as that in the 2-STAP scheme, but in the second stage we make measurements on mixtures (groups) of positive pools together, instead of making measurements on separate pools only. For the ease of exposition, we explain a special case of the 2-STAMP scheme when up to two pools can be mixed together. The same idea can be easily extended for mixing larger number of pools.

The main idea behind mixing pools in the second stage is as follows. Consider two positive pools that we expect to contain a relatively small number of infected people. (For each positive pool, the number of infected people can be estimated based on the observed measurement for that pool in the first stage, for details, see [12]). By mixing these pools together and making measurements on the mixed pool altogether (instead of making measurements on the pools individually), we can save a few measurements while maintaining the implementation/computational complexity of both the pooling and recovery algorithms affordable. However, the rest of the pools that are expected to contain a relatively large number of nonzero coordinates will be measured individually, so as to avoid the pooling and/or recovery algorithms to become too complex implementation-wise or computationally. Details about the mixing process and pooling matrices we have used in our Monte-Carlo simulations can be found in Appendix II, and the detailed description of the recovery algorithm can be found in [12].

## V. Comparisons Between the Proposed Schemes and the Tapestry Scheme

The key differences between the proposed schemes and the Tapestry scheme are listed below:

- Tapestry is a single-stage scheme, and hence all measurements can be made in parallel. The proposed schemes are, however, two-stage schemes; and notwithstanding that all measurements in each stage can be made in parallel, the measurements in the second stage can only be made after those in the first stage.
- When compared to Tapestry, in the tested cases, the proposed schemes achieve a better trade-off between the average number of measurements and the (conditional) average false negative/positive rate. This comes from two facts: (i) the pooling algorithm of the proposed schemes is more flexible than that of the Tapestry scheme. This is because the total number of measurements in the latter can vary for different realizations of viral loads, whereas the former uses the same number of measurements always; and (ii) the measurements in the proposed schemes are localized to small pools. This makes it possible to implement recovery algorithms that are carefully designed for the multiplicative noise model with reasonable computational complexity. In contrast, the recovery algorithms discussed in the Tapestry scheme were borrowed from the compressed sensing literature where the noise model is assumed to be additive. Unlike the recovery algorithms used for Tapestry, the recovery algorithm proposed in this work also takes into account the signal and noise distributions and the sparsity parameter (prevalence) or its estimate.
- The pooling algorithm of the proposed schemes can potentially have a substantially lower computational and implementation complexity than that of the Tapestry scheme. This follows from two facts: (i) the total number of nonzero entries in the overall pooling matrix of the proposed schemes can be much smaller than that in the Tapestry scheme; and (ii) the nonzero entries in each row of the pooling matrix in the Tapestry scheme are spread out everywhere, whereas the nonzero entries in the pooling matrix of the proposed schemes are localized in each row. In particular, in the first stage each measurement is localized to a pool of people with consecutive labels, and the measurements in the second stage for each pool are over the people in that pool only.

## VI. Monte-Carlo Simulation Results

In this section, we present our simulation results. As a case study, in these simulations, we have considered a population of *n* = 961 people to be tested for COVID-19 and assumed that the prevalence is *p* = 0.01. (In the simulations, we considered both cases where *p* is known or unknown *a priori*, and we did not observe any significant difference in the performance of either of the proposed schemes, 2-STAP and 2-STAMP.) For both the proposed schemes, we have considered pooling the population into *q* = 31 pools, each of size 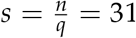, in the first stage. Three different values of number of infected people in the population (*k*), namely *k* ∈ *{*5, 10, 15*}*, have been considered. For every *k*, we performed 100 Monte-Carlo simulations, where the statistical models used for viral load and measurement noise were obtained from [11].

Table I summarizes our results for the proposed 2-STAP (both variants) and 2-STAMP schemes and the results for the Tapestry scheme for the same problem model (i.e., the same population size and the same viral load and noise distributions) where each measurement is made on a pool of 31 people (see [11]). In this table, *m*_min_, *m*_max_, *m*_std_, and *m*_ave_ represent the minimum, maximum, standard deviation, and the average of the number of measurements used in 100 simulations, respectively. The sensitivity and specificity results are rounded to two decimal places, for fair comparison with the results reported in [11]. More detailed results for the 2-STAP-I, 2-STAP-II, and 2-STAMP schemes are presented in Tables II, III, and IV, respectively. In these tables, *α* is a parameter used in the recovery algorithm that controls the trade-off between sensitivity and specificity (for more details, see [12]).

**TABLE I.** Performance results for the Tapestry scheme [11] and the proposed 2-stap and 2-STAMPschemes (sensitivity and specificity results are rounded to two decimal places)

| $k$ | $m_{\min}$ | $m_{\max}$ | $m_{\text{std}}$ | $m_{\text{ave}}$ | Pooling Scheme | Conditional Sensitivity | Conditional Specificity |
| --- | --- | --- | --- | --- | --- | --- | --- |
| 5 | 93 | 93 | 0 | 93 | Tapestry + COMP + NN-LASSO [11] | 1.00 | 1.00 |
|  |  |  |  |  | Tapestry + COMP + NN-OMP [11] | 1.00 | 1.00 |
|  |  |  |  |  | Tapestry + COMP + SBL [11] | 1.00 | 1.00 |
|  | 49 | 61 | 2.94 | 59.08 | 2-STAP-I | 1.00 | 1.00 |
|  | 47 | 57 | 2.26 | 54.55 | 2-STAP-II | 1.00 | 1.00 |
|  | 46 | 55 | 2.25 | 52.56 | 2-STAMP | 1.00 | 1.00 |
| 10 | 93 | 93 | 0 | 93 | Tapestry + COMP + NN-LASSO [11] | 0.98 | 0.99 |
|  |  |  |  |  | Tapestry + COMP + NN-OMP [11] | 0.96 | 1.00 |
|  |  |  |  |  | Tapestry + COMP + SBL [11] | 0.99 | 0.99 |
|  | 73 | 91 | 5.54 | 82.96 | 2-STAP-I | 1.00 | 1.00 |
|  | 66 | 81 | 4.18 | 74.98 | 2-STAP-II | 0.99 | 1.00 |
|  | 63 | 76 | 3.74 | 70.86 | 2-STAMP | 1.00 | 1.00 |
| 15 | 93 | 93 | 0 | 93 | Tapestry + COMP + NN-LASSO [11] | 0.94 | 0.97 |
|  |  |  |  |  | Tapestry + COMP + NN-OMP [11] | 0.86 | 0.99 |
|  |  |  |  |  | Tapestry + COMP + SBL [11] | 0.98 | 0.97 |
|  | 85 | 121 | 7.31 | 103.30 | 2-STAP-I | 0.98 | 0.99 |
|  | 78 | 106 | 5.65 | 92.66 | 2-STAP-II | 0.98 | 0.99 |
|  | 74 | 99 | 5.02 | 86.85 | 2-STAMP | 0.99 | 0.99 |

**TABLE II.** Detailed performance results for the 2-STAP-I scheme

| $k$ | $m_{\min}$ | $m_{\max}$ | $m_{\text{std}}$ | $m_{\text{ave}}$ | $\alpha$ | Conditional Sensitivity | Conditional Specificity |
| --- | --- | --- | --- | --- | --- | --- | --- |
| 5 | 49 | 61 | 2.94 | 59.08 | 0.943 | 0.9920 | 0.9995 |
|  |  |  |  |  | 0.926 | 0.9960 | 0.9994 |
|  |  |  |  |  | 0.893 | 0.9980 | 0.9993 |
| 10 | 73 | 91 | 5.54 | 82.96 | 0.943 | 0.9810 | 0.9988 |
|  |  |  |  |  | 0.909 | 0.9930 | 0.9982 |
|  |  |  |  |  | 0.885 | 0.9950 | 0.9978 |
| 15 | 85 | 121 | 7.30 | 103.30 | 0.847 | 0.9820 | 0.9939 |
|  |  |  |  |  | 0.758 | 0.9893 | 0.9885 |
|  |  |  |  |  | 0.649 | 0.9940 | 0.9817 |

**TABLE III.** Detailed performance results for the 2-STAP-II scheme

| $k$ | $m_{\min}$ | $m_{\max}$ | $m_{\text{std}}$ | $m_{\text{ave}}$ | $\alpha$ | Conditional Sensitivity | Conditional Specificity |
| --- | --- | --- | --- | --- | --- | --- | --- |
| 5 | 47 | 57 | 2.26 | 54.55 | 0.813 | 0.9920 | 0.9986 |
|  |  |  |  |  | 0.794 | 0.9940 | 0.9983 |
|  |  |  |  |  | 0.787 | 0.9980 | 0.9982 |
| 10 | 66 | 81 | 4.18 | 74.98 | 0.943 | 0.9810 | 0.9987 |
|  |  |  |  |  | 0.794 | 0.9930 | 0.9954 |
|  |  |  |  |  | 0.730 | 0.9950 | 0.9927 |
| 15 | 78 | 106 | 5.65 | 92.66 | 0.787 | 0.9820 | 0.9903 |
|  |  |  |  |  | 0.654 | 0.9893 | 0.9802 |
|  |  |  |  |  | 0.515 | 0.9940 | 0.9724 |

**TABLE IV.**
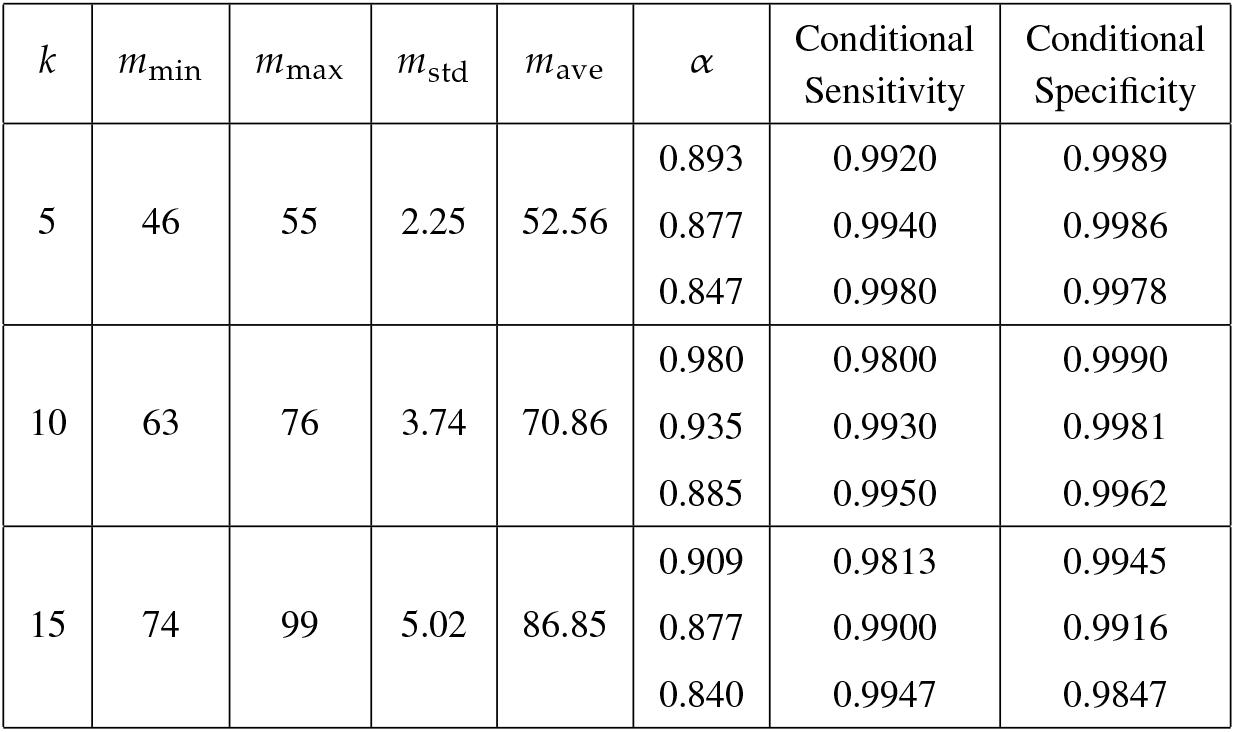
Detailed performance results for the 2-STAMP scheme

Comparing the results of Tapestry and the two variants of 2-STAP in Table I, it can be seen that for *k* ∈ *{*5, 10*}*, 2-STAP-I requires smaller number of measurements on the average for the same (or even higher) sensitivity and specificity (see also Table II). For *k* = 15, 2-STAP-I uses about 10 more measurements than Tapestry on the average, but it achieves a substantially higher specificity by about 2% for almost the same sensitivity (see also Table II). It can also be seen that for all *k* ∈ *{*5, 10, 15*}*, 2-STAP-II can provide higher sensitivity and higher specificity than Tapestry with even smaller (average) number of measurements. For instance, for the case of *k* = 10 infected people, with an average number of measurements about 75, 2-STAP-II can achieve a sensitivity of 99.30% and a specificity of 99.54%, see Table III. However, using Tapestry, for *k* = 10, one can achieve a sensitivity and a specificity between 98.50% and 99.49% with 93 measurements (about 20% more measurements than that in 2-STAP-II). These improvements in the performance are mainly due to the fact that 2-STAP is an adaptive scheme (although with a very small degree of adaptivity, i.e., using only one round of feedback), whereas Tapestry is a non-adaptive scheme. In particular, identifying all negative pools (which contain a relatively large fraction of population for sufficiently low prevalence) at the end of the first stage and using a relatively small number of additional measurements only for each positive pool in the second stage enable us to achieve a better trade-off between average number of measurements, sensitivity, and specificity.

As can be seen in Table I, 2-STAMP can achieve a sensitivity and a specificity higher than those attainable with 2-STAP-I and 2-STAP-II, with even smaller average number of measurements. For instance, for *k* = 10, with only about 71 (< 75 in 2-STAP-II) measurements on average, 2-STAMP can achieve a sensitivity of 99.30% (the same as that in 2-STAP-II) and a specificity of 99.81% (> 99.54% in 2-STAP-II), see Table III. The advantage of 2-STAMP over both variants of 2-STAP comes from the saving in the number of measurements in the second stage. In particular, in 2-STAMP, mixing small groups (namely, groups of size two) of pools with small number of infected people gives rise to an opportunity for making a smaller number of measurements on the mixed super-pool (as compared to the total number of measurements used in 2-STAP for all pools in the mix) without compensating the overall accuracy.

## Data Availability

No clinical data were used.

## Appendix I

### Detailed Explanation about Multiplicative Noise Model

The multiplicative noise model is inspired by the current RT-qPCR technology for COVID-19 testing. Suppose that an individual person is to be tested for COVID-19 by using this technology. The sample collected from the person to be tested is dispersed into a liquid medium, and the reverse transcription (RT) process is applied to convert the RNA molecules of the SARS-CoV-2 virus (the coronavirus that causes COVID-19) in the liquid (if the person is infected) into cDNA. Followed by adding primers that are complementary to the cDNA of the viral genome, these primers attach themselves to the cDNA of the viral genome, and together they undergo an exponential amplification process by the RT-qPCR machine [11]. This process consists of a maximum of *C*_max_ cycles. The output of the RT-qPCR process is the cycle count *C* after which the concentration of DNA exceeds a pre-specified threshold *D*_min_ or *C*_max_. The thresholds *D*_min_ and *C*_max_ are often chosen so that: (i) if a person is not infected, the DNA concentration does not exceed *D*_min_ over the course of *C*_max_ cycles, and (ii) if a person is infected, the DNA concentration exceeds *D*_min_ at some point, say cycle *C*, over the course of *C*_max_ cycles. Note that for a fixed *D*_min_, the larger is the viral load of an infected person, the smaller *C* will be. Ideally, the concentration of DNA molecules is doubled in every cycle, i.e., after *C* cycles the concentration of DNA molecules is *x*2^*C*^, where *x* is the viral load of the person to be tested. In reality, however, the amplification process may not be ideal. To reflect the randomness in the process, we use the same model as the one suggested in [11] and assume that the concentration of DNA molecules after *C* cycles is given by *xb*^*C+*Δ^ for some positive constant *b* (close to 2), where *x* is the viral load of the person to be tested, and Δ is a Gaussian random variable with mean zero and variance 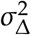. The multiplicative term *ε* = *b*^Δ^ can be viewed as the noise in the amplification process. Note that the closer are *b* to 2 and *σ*_Δ_ to 0, the weaker will be the noise and the closer will be the process to ideal. For any *x* > 0, let *C*_*x*_ be the number of cycles it takes for the concentration of DNA molecules to be approximately equal to *D*_min_. That is,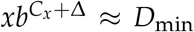. The cycle count measurement *C*_*x*_ can be converted to an equivalent measurement *z* given by 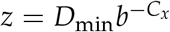. It can then be seen that *z* = *xε* gives the measurement model with multiplicative noise, as in (1).

## Appendix II

### Number of Measurements and Pooling Matrices Used in Simulations

For the 2-STAP-I scheme, for each positive pool, 6 measurements were used in the second stage, and the pooling matrix used in the simulations can be found in Fig. 3.

**Fig. 3.**
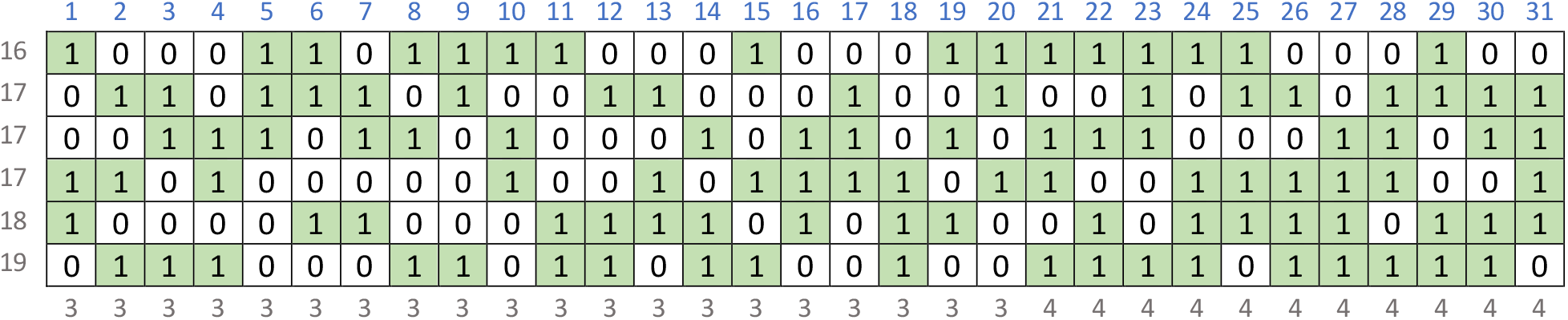
The pooling matrix with 6 rows and 31 columns used in the simulations for the 2-STAP-I scheme. (The number of 1’s in each row is shown in front of the row, and the number of 1’s in each column is shown below the column.)

For the 2-STAP-II scheme, the number of measurements used in the second stage for each positive pool was chosen as follows: 5, 6, 7, 8 measurements if our estimate of the number of infected people in the pool (based on the observed measurement in the first stage for that pool) is 1, 2, 3, ≥ 4, respectively. The pooling matrices used in the simulations for these number of measurements can be found in Fig. 4.

**Fig. 4.**
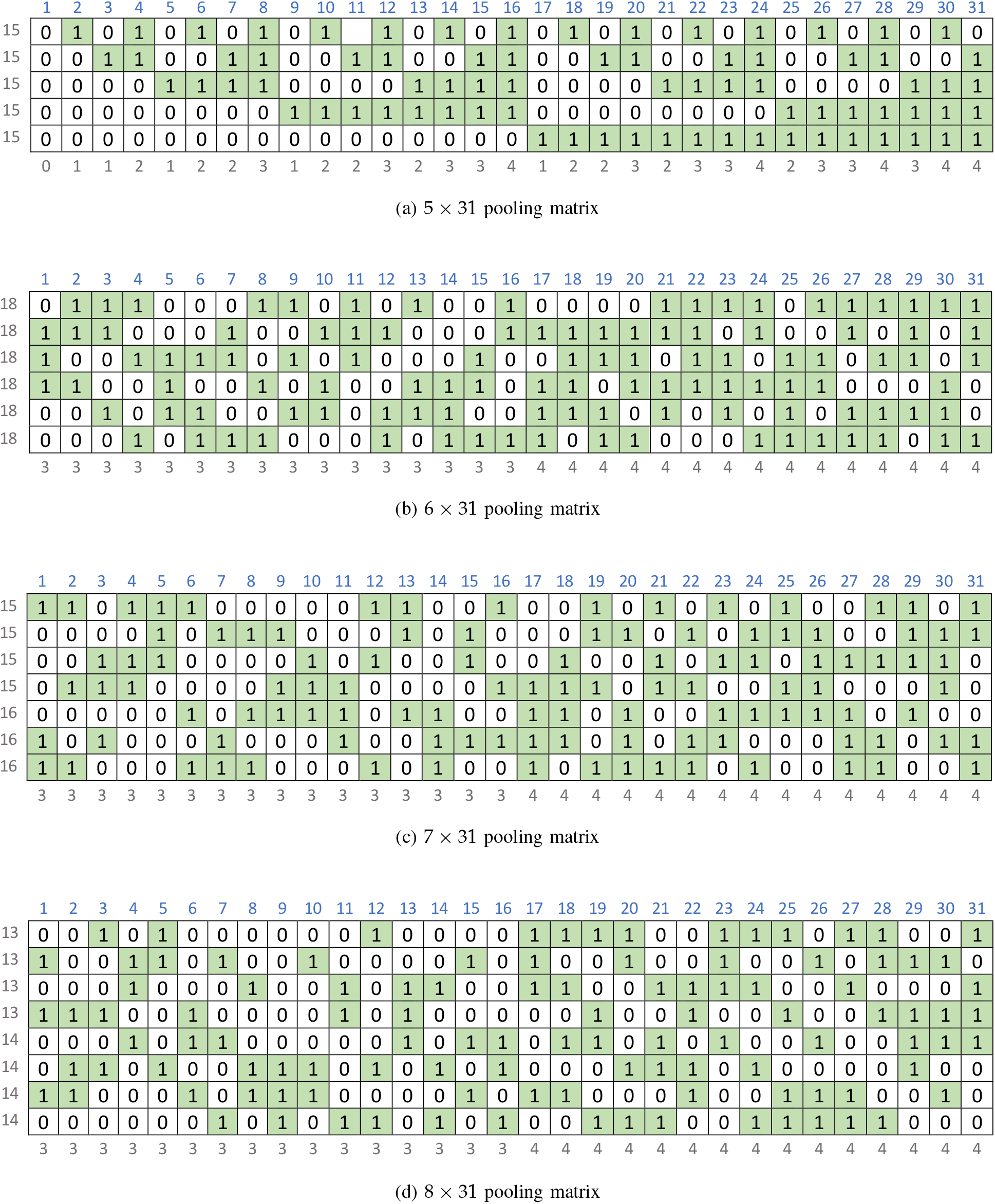
The pooling matrices with 5, 6, 7,8 rows and 31 columns used in the simulations for the 2-STAP-II scheme. (The number of 1’s in each row is shown in front of the row, and the number of 1’s in each column is shown below the column.)

For the 2-STAMP scheme, we first estimated the number of infected people in each positive pool based on the observed measurement for that pool in the first stage. Then, we sorted the pools in the decreasing order with respect to their estimated number of infected people. For any pool for which the (estimated) number of infected people is 3 or ≥ 4, we used 7 or 8 measurements, respectively. The pooling matrices used in the simulations for these number of measurements can be found in Fig. 4.

For any two consecutive pools (in the sorted order) that collectively contain 2, 3, 4 infected people (based on our estimates), we used 9, 10, 11 measurements, respectively, on the mixed pool formed by combining the two pools together. The pooling matrices used in the simulations for these number of measurements can be found in Fig. 5.

**Fig. 5.**
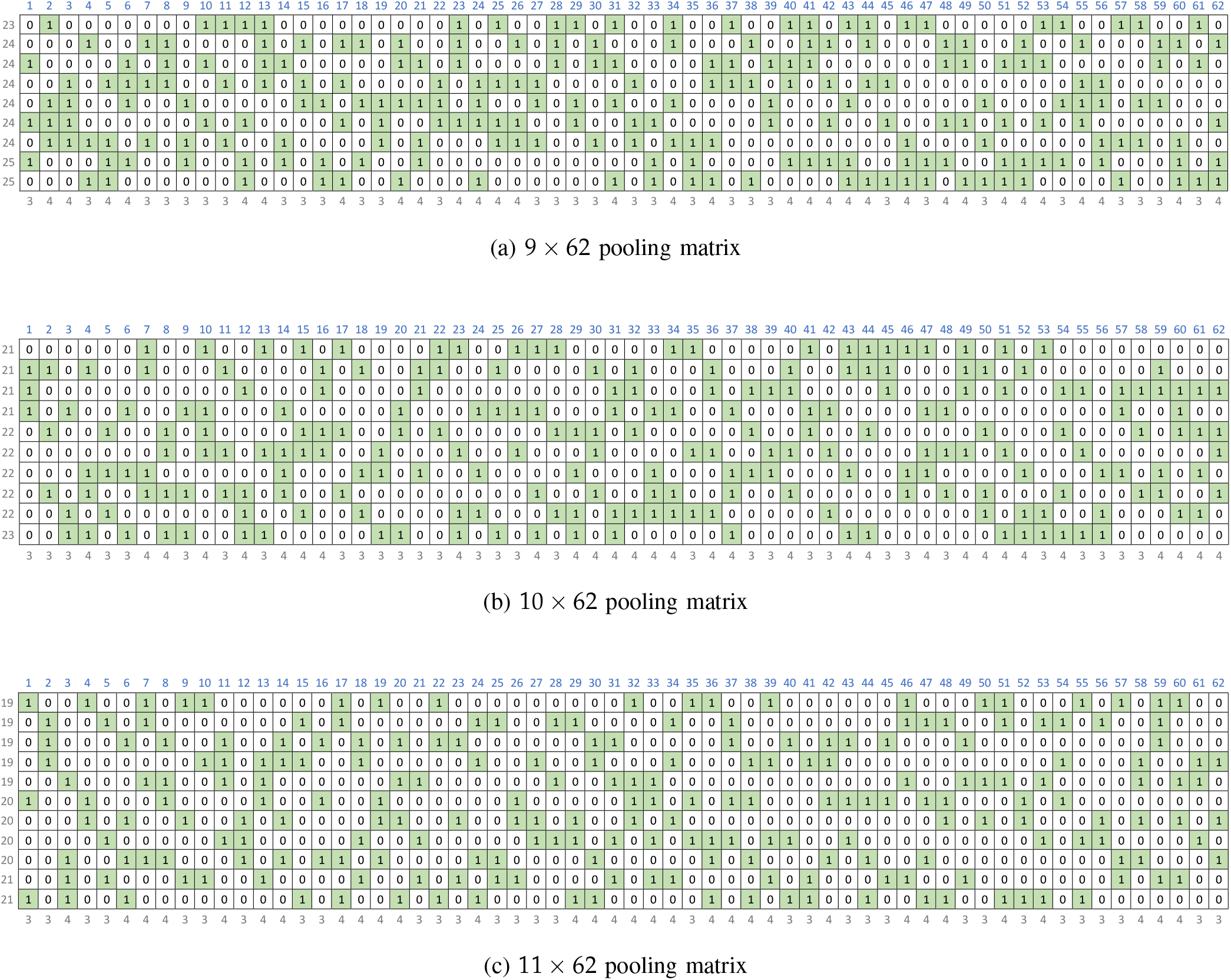
The pooling matrices with 9, 10, 11 rows and 62 columns used in the simulations for the 2-STAMP scheme. (The number of 1’s in each row is shown in front of the row, and the number of 1’s in each column is shown below the column.)

